# An Online Risk Calculator for Rapid Prediction of In-hospital Mortality from COVID-19 Infection

**DOI:** 10.1101/2021.01.22.21249953

**Authors:** Daniel S. Evans, Kyoung Min Kim, Xiaqing Jiang, Jessica Jacobson, Warren Browner, Steven R. Cummings

## Abstract

Prediction of mortality from COVID-19 infection might help triage patients to hospitalization and intensive care. To estimate the risk of inpatient mortality, we analyzed the data of 13,190 adult patients in the New York City Health + Hospitals system admitted for COVID-19 infection from March 1 to June 30, 2020. They had a mean age 58 years, 40% were Latinx, 29% Black, 9% White and 22% of other races/ethnicities and 2,875 died. We used Machine learning (Gradient Boosted Decision Trees; XGBoost) to select predictors of inpatient mortality from demographics, vital signs and lab tests results from initial encounters. XGBoost identified O_2_ saturation, systolic and diastolic blood pressure, pulse rate, respiratory rate, age, and BUN with an Area Under the Receiver Operating Characteristics Curve = 94%. We applied CART to find cut-points in these variables, logistic regression to calculate odds-ratios for those categories, and assigned points to the categories to develop a score. A score = 0 indicates a 0.8% (95% confidence interval, 0.5 – 1.0%) risk of dying and ≥ 12 points indicates a 98% (97-99%) risk, and other scores have intermediate risks. We translated the models into an online calculator for the probability of mortality with 95% confidence intervals (as pictured):

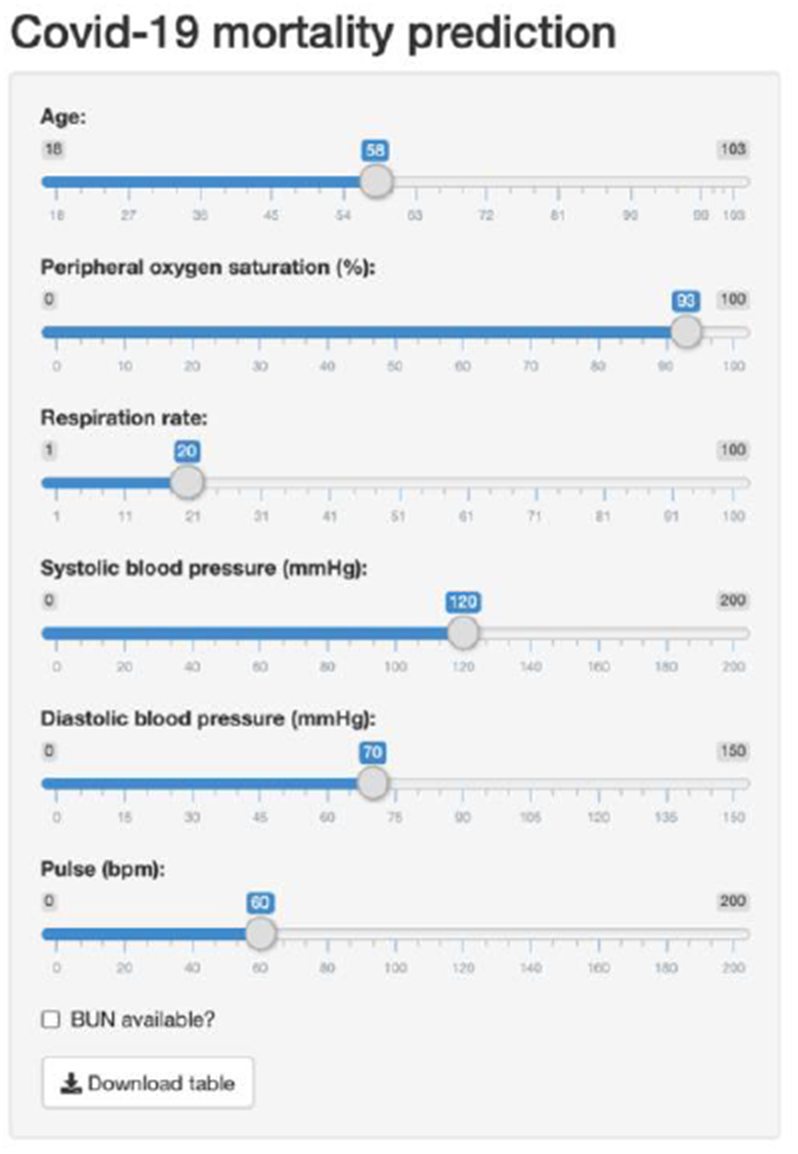

danielevanslab.shinyapps.io/COVID_mortality/

---

Hospitals have been overwhelmed by admissions of patients with COVID 19 infection. Accurate prediction of the probability of death might help prioritize patients for hospitalization and intensive care. Predictive models would be useful if they are available in a form, such as an online calculator, that a clinician could readily use in a busy acute care setting. No previous study has generated models from large diverse populations and based on assessments readily available when decisions are made about hospitalization and ICU admission.

## Methods

We used data from all patients ≥ 18 years old who admitted to New York City Health + Hospitals (NYC H+H) system between March 1 and June 30, 2020. The outcome was death from any cause at any time during the hospitalizations. We selected demographic characteristics (sex, age, race and ethnicity), weight, BMI, vital signs, O2 saturation (SpO2) from peripheral oxygen saturation monitors, and routinely ordered clinical laboratory tests.

The goal was to develop an online calculator based on a model to estimate the probability of dying in hospital for patients admitted for COVID-19 infection. We used Extreme Gradient Boosted Decision Trees (XGBoost), followed by the identification of cut-points of the selected variables using classification and regression trees (CART), then developed a score to predict mortality. Train and test data partitions were created using an 80% and 20% random split stratified by death status to ensure an even proportion of mortality in the train and test datasets. Gradient Boosted Decision Trees implemented in the XGBoost R package v 1.2.0.1 with R v 4.0.2. was used to generate an ensemble of multiple decision trees to minimize errors in the classification of mortality in patients. The XGBoost model was developed in the train partition, using four boosting rounds, a maximum depth of three for each decision tree, a learning rate of 0.3, a binary:logistic learning objective with error rate used as the evaluation metric with a minimum child weight of 75. Feature importance was evaluated using the information gain metric of a split on a variable. XGBoost model performance was evaluated in the test partition using accuracy and area under the curve (AUC) from a receiver operating characteristic (ROC) curve. Selected features and model performance did not change with 10-fold cross-validation.

To develop a clinical prediction score, in the original 80% training set, we analyzed each variable by Classification and Regression Trees (CART) to identify optimum cut-points for classifying patients as died or survived. We applied logistic regression to the categorical variables to develop a multivariate model predicting death. The coefficients for each value of the variable were assigned a point value by rounding its coefficient to the nearest integer. The proportion of patients who died was calculated for each 1-point interval. The highest risk categories with very similar risk score and small number of patients were combined. This analysis was repeated in the original 20% test set to assess the consistency of the clinical prediction score and estimated probabilities of dying. AUC value from a ROC curve was evaluated to test the performance of clinical prediction score.

To develop a clinical prediction score, we used Classification and Regression Tree (CART) analyses in the original 80% training set to identify optimum cut-points for each variable selected by XGBoost. There was no cut-point for creatinine and it had low importance in the XGBoost model, therefore it was not included in the final calculation of clinical risk score. There were 1,501 patients missing a BUN value. To determine whether a missing value was associated with risk of mortality, we created an indicator variable for a missing BUN value. We entered the selected variables and cut-points into a logistic regression model to estimate the multivariable odds ratios. To assign risk scores, the odds ratio for each of these categorical variables were divided by 2.6 (the lowest odds ratio), rounded, and then summed for each patient to calculate a risk score. After excluding 703 patients with missing values for one or more variables, the proportions of patients who died were calculated for each 1-point interval in risk score; the highest-risk categories, which had similar scores and small numbers of patients, were combined. Because the predicted mortality by risk score categories were very similar in the training and test sets, these sets were combined to estimate the probabilities and 95% confidence intervals for the entire population.

## Results

The cohort included 5421 women, had a mean age 58 years, 5258 were Latinx [40%], 3805 Black [29%], 1168 White [9%], and 2959 individuals of other races/ethnicities [22%] (Table 1). A total of 2,875 (21.8%) died. The XGBoost algorithm identified eight variables (Figure 1) that, together predicted mortality with an AUC = 94% and an accuracy of 91% (Figure 2). The four boosted decision trees are provided Figure 3 a-d. Of the variables that the XGBoost analysis selected for the algorithm, SpO2 was the strongest predictor. Respiratory rate, blood pressures, pulse rate and age were also important predictors. A multivariable logistic model showed that the selected cut-points were all significant predictors of mortality (Table 1). The risk score based on the odds ratios for these variables ranged from 0 to 22 points and had an AUC of 0.94 for predicting mortality (Figure 2). There were 5,677 (45.5%) patients with a score of 0, and 674 (5.4%) with a score ≥ 12 points (Table 2). In-hospital mortality ranged from 0.8% (95% confidence interval, 0.5 - 1.0 %) for those with a score of 0 to 97.6% (96.5 - 98.8%) for patients with a score ≥ 12 points.

**Table 1.** Predictors from the Multivariable Model and Points Indicating an Increased Odds of In-hospital Death

|  | *Odds ratio*<br>*(95% CI)* | *Points* |
| --- | --- | --- |
| *Age* |  |  |
| Age < 70 years old | Reference | 0 |
| 70 ≤ Age < 85 years old | 2.6 (2.2-3.1) | 1 |
| Age ≥ 85 years old | 5.4 (4.2-7.0) | 2 |
| *O2 Saturation (SpO2)* |  |  |
| SpO2 ≥ 91% | Reference | 0 |
| SpO2 < 91% | 10.7 (8.3-19.9) | 4 |
| *Respiratory Rate (RR)* |  |  |
| 14 ≤ RR < 22/min | Reference | 0 |
| RR ≥ 22/min | 7.8 (4.2-14.4) | 3 |
| RR < 14/min | 9.2 (7.6-11.1) | 4 |
| *Pulse Rate (PR)* |  |  |
| 51 ≤ PR < 109/min | Reference | 0 |
| PR < 51/min | 3.5 (2.6-4.7) | 1 |
| 109 ≤ PR < 119/min | 9.7 (4.8-20.0) | 4 |
| PR ≥ 119/min | 12.5 (9.1-17.3) | 5 |
| *Systolic BP (SBP)* |  |  |
| SBP ≥ 95 mmHg | Reference | 0 |
| SBP < 95 mmHg | 7.9 (5.8-10.7) | 3 |
| *Diastolic BP (DBP)* |  |  |
| DBP ≥ 54 mmHg | Reference | 0 |
| DBP < 54 mmHg | 4.7 (3.6-6.1) | 2 |
| *BUN* |  |  |
| BUN < 20 mmHg | Reference | 0 |
| 20 ≤ BUN < 44 mmHg | 2.6 (2.1-3.2) | 1 |
| BUN ≥ 44 mmHg | 5.9 (4.8-7.4) | 2 |
| If BUN value is missing | 0.2 (0.1-0.4) | 0 |
| Range of risk score |  | 0-22 |

BP, blood pressure; BUN, blood urea nitrogen

**Table 2.** Total Point Score and Risk of In-hospital Death

| Total<br>Score | Risk of Death %<br>(95% C.I.) |
| --- | --- |
| 0 | 0.8 (0.5-1.0) |
| 1 | 4.5 (3.5-5.5) |
| 3 | 9.7 (8.0-11.4) |
| 4 | 20.7 (17.6-23.8) |
| 5 | 39.4 (34.6-44.1) |
| 6 | 57.8 (51.9-63.8) |
| 7 | 64.3 (58.6-69.9) |
| 8 | 74.4 (69.2-79.7) |
| 9 | 87.6 (82.8-92.4) |
| 10 | 92.3 (88.6-95.9) |
| 11 | 92.4 (88.2-96.6) |
| ≥12 | 97.6 (96.5-98.8) |

**Figure 1.**
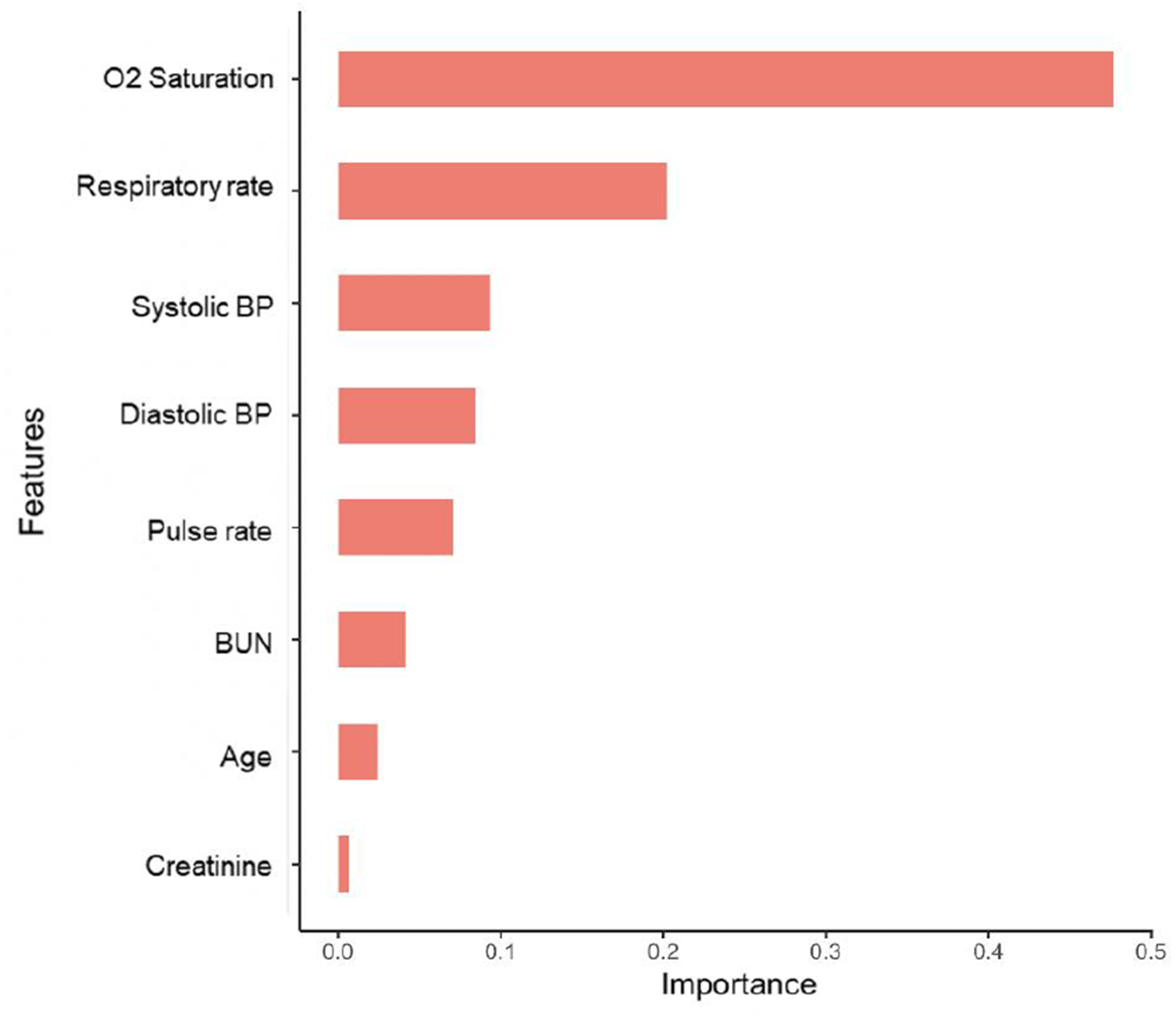
Features included in the XGBoost Model Ranked by Gain in the Accuracy of Classification when Used in Decision Trees that Generated the Model

**Figure 2.**
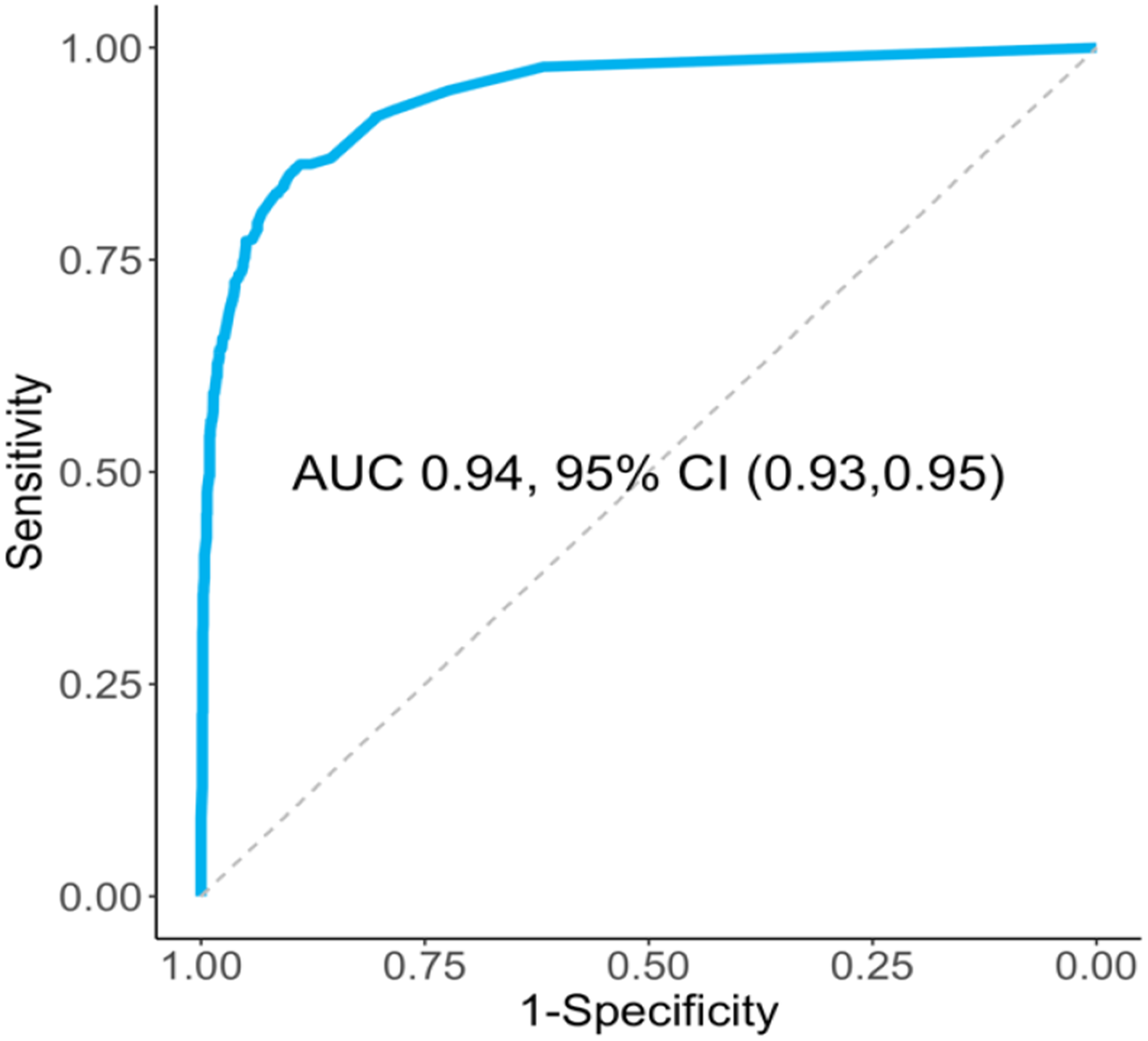
Receiver Operating Characteristics (ROC) Curves of Mortality Predicted by the Machine Learning XGBoost Model

**Figure 3.**
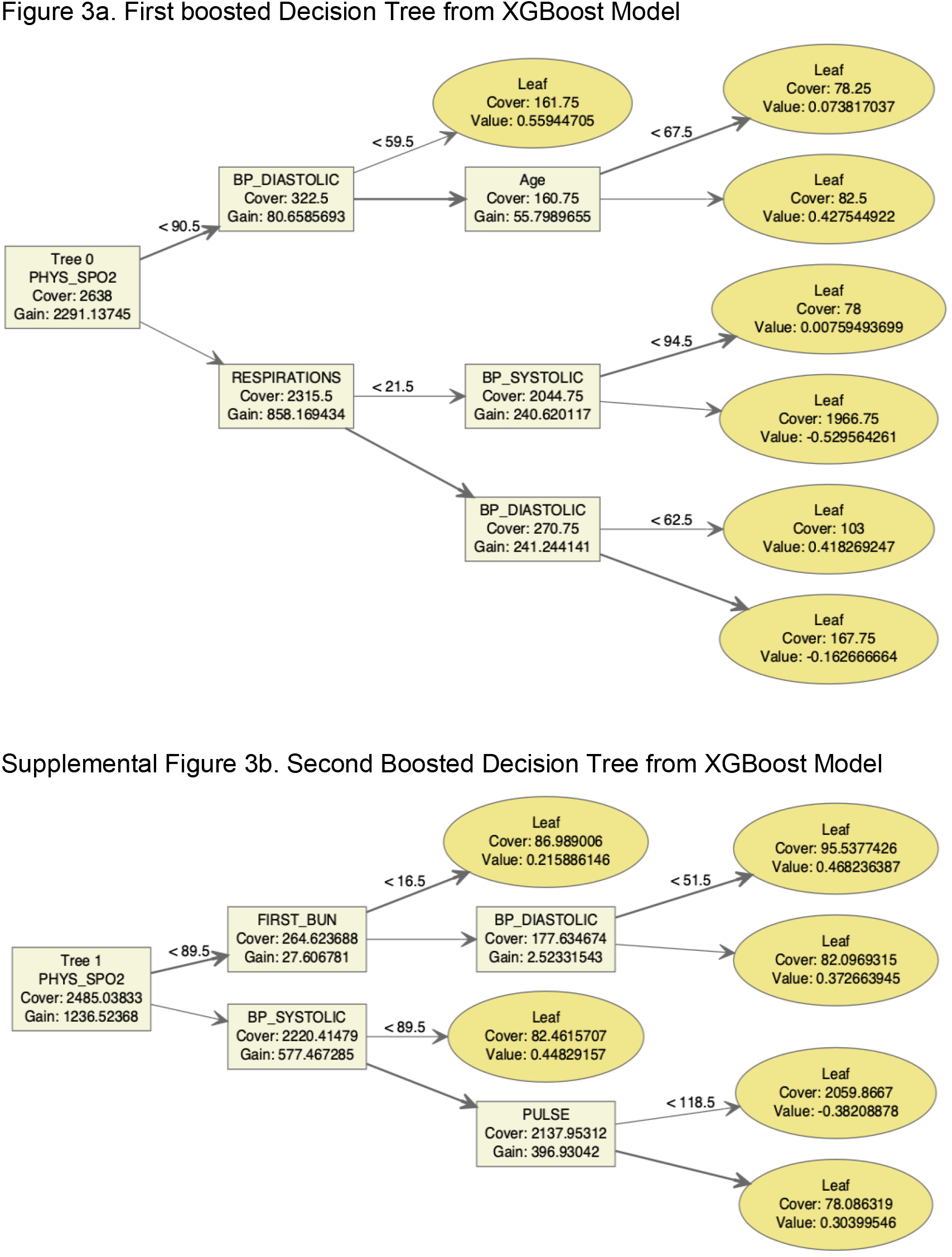
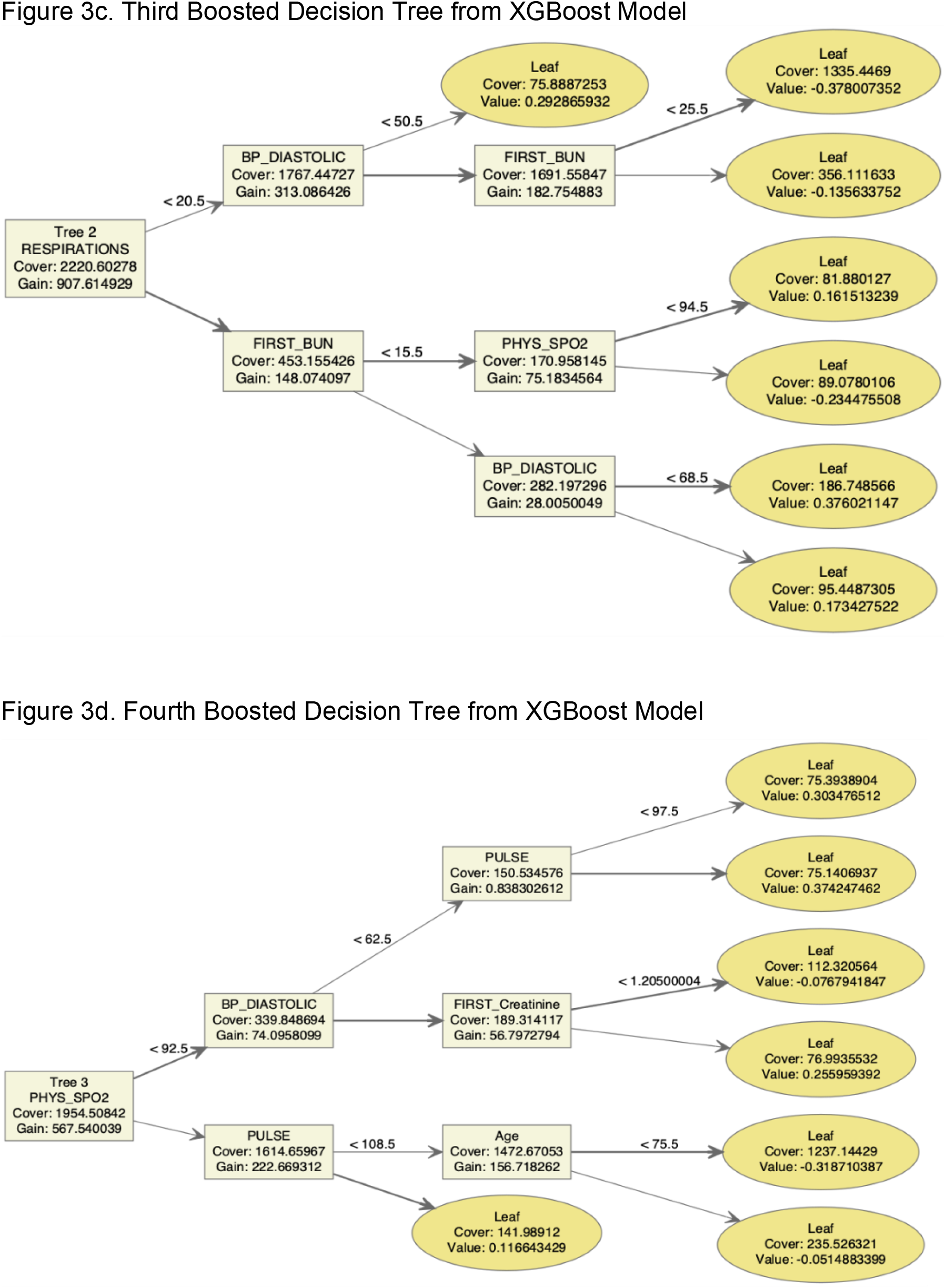
Boosted Decision Trees from the XGBoost Models. Cover is the sum of the second order gradient of training data classified to the leaf. Gain is the information gain metric of a split. Value is the margin value that the leaf contributes to prediction.

**Figure 4.**
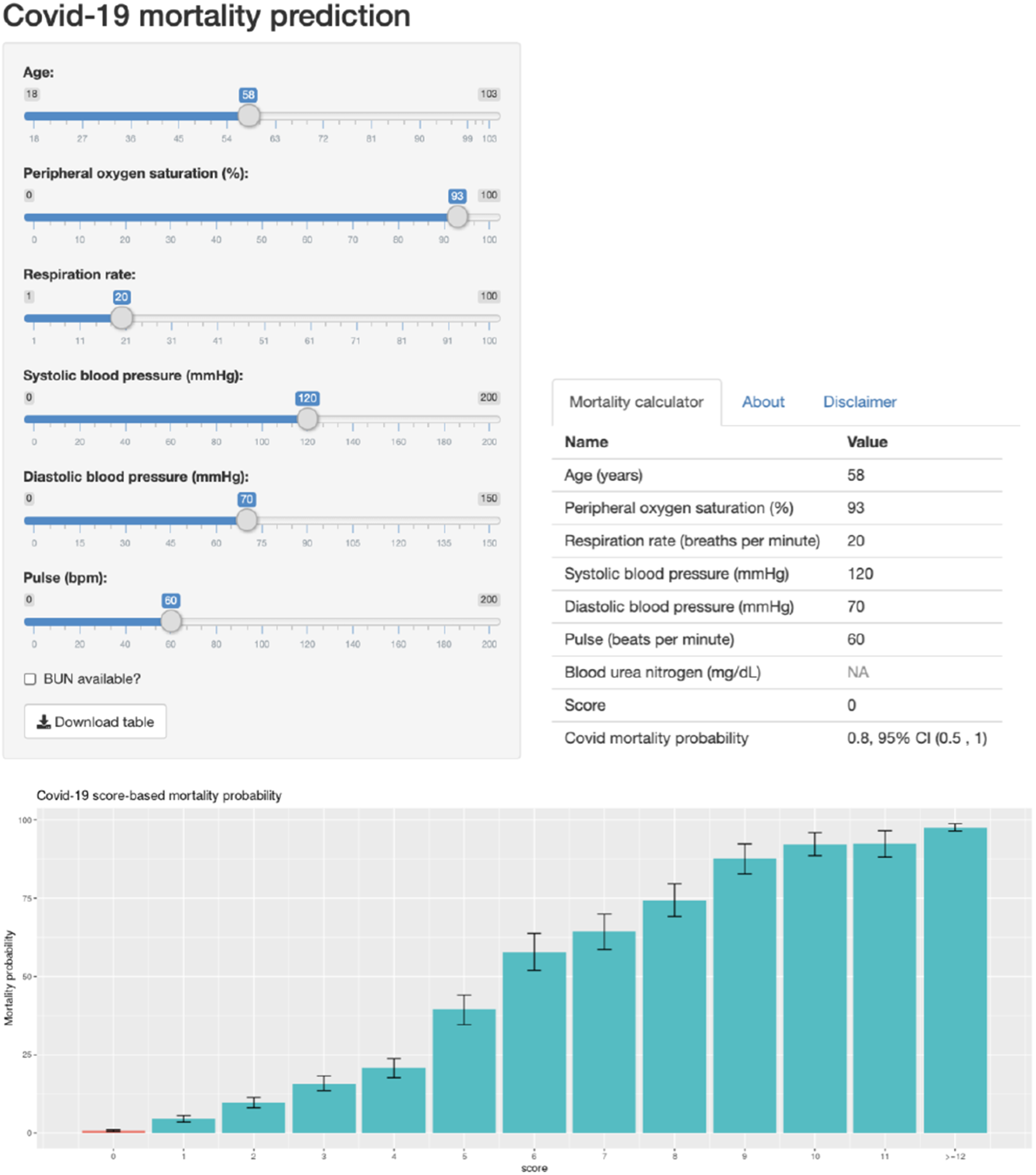
Screen shot of Covid-19 calculator at https://danielevanslab.shinyapps.io/Covid_mortality/

The online calculator reports the probability of in-hospital mortality based on the risk score (danielevanslab.shinyapps.io/Covid_mortality). To report the probability of dying, all variables must have non-missing values except for the blood urea nitrogen (BUN) test which includes a term for missing: https://danielevanslab.shinyapps.io/Covid_mortality/.

## Discussion

We found that a few variables available in the initial assessment of patients with COVID-19 infection can estimate the probability of dying during hospitalization. The model that estimates the probability of dying is available online for convenient use in acute care settings. SpO2 was the strongest predictor and vital signs were also important. BUN was the only laboratory value in the algorithm but it is optional for calculation of the risk of death. The data were collected before effective treatments, such as corticosteroids, were commonly used in the treatment of Covid 19. Therefore, the model identifies the probability of dying without current in-hospital treatments and, therefore, may identify patients who are most likely to benefit from hospital care.

This analysis has strengths. The algorithm is derived from the very diverse population in New York City and, therefore, may be generalizable to patients of all races and ethnicities. The study population was very large which produced estimates of mortality that have very narrow confidence limits. The algorithm produced consistent results by 10-fold cross-validation and testing its performance in a random 20% of the data. A limitation of the model is that the data did not include measurements, such as markers of inflammation and coagulation, comorbidities, and indices of comorbidity and severity of illness that have been demonstrated in other studies to predict mortality.

## Data Availability

The patient data are not available. Access to the data was granted for the sole purpose of this manuscript.

https://danielevanslab.shinyapps.io/Covid_mortality/

## Acknowledgements

The authors have no financial conflicts of interest.

